# Establishment of human post-vaccination SARS-CoV-2 standard reference sera

**DOI:** 10.1101/2022.01.24.22269773

**Authors:** Jinhua Xiang, Louis Katz, Patricia L. Winokur, Ashok Chaudhary, Rebecca Bradford, Sujatha Rashid, Sudakshina Ghosh, Angela Robertson, Joseph Menetski, Taylor Lee, Brittany Poelaert, Richard T. Eastman, Matthew D. Hall, Jack T. Stapleton

## Abstract

As SARS-CoV-2 variants emerge, there is a critical need to understand the effectiveness of serum elicited by different SARS-CoV-2 vaccines. A reference reagent comprised of post-vaccination sera from recipients of different vaccines allows evaluation of *in vitro* variant neutralization, and provides a reference for comparing assay results across laboratories. We prepared and pooled >1 L serum from donors who received the SARS-CoV-2 mRNA vaccines (BNT162b2, Pfizer and mRNA-1273, Moderna), a replication-incompetent adenovirus type 26 vaccine (Ad26.COV2.S, Johnson and Johnson), or recombinant spike protein expressed by baculovirus incorporated into a nanoparticle vaccine plus Matrix-M adjuvant (NVX-CoV2373, Novavax). Twice frozen sera were aliquoted and are available for distribution to the research community (BEI Resources). The calculated WHO titer of pooled sera to spike protein was 1,312, 1,447, 1,936, and 587 and the reciprocal RBD binding to ACE-2 IC90-titers were 60, 64, 118, and 46 for BNT162b2, mRNA1273, Ad26.CoV2373, and NVX-CoV2373 sera, respectively.

## INTRODUCTION

RNA viruses, including the severe acute respiratory syndrome coronavirus 2 (SARS-CoV-2), have error-prone polymerases that result in high rates of genome mutation (1, 2). This results in progeny virus genomes that differ from their parental templates, leading to the generation of viral quasispecies within and between individuals (3). SARS-CoV-2 has evolved into variants of concern (VOC) that demonstrate mutations, including the gene that encodes the spike protein that mediates viral entry. These mutations may lead to enhanced transmissibility and new waves of infections (4), as seen most recently Delta (B.1.617.2 and sub-lineages) and Omicron (B.1.1.529) VOC (5, 6). Among VOC, a critical issue is their potential for altered pathogenicity and immune escape from naturally occurring or vaccine-induced antibodies (7).

As new SARS-CoV-2 variants emerge, there is an urgent need to understand the relative effectiveness of antibodies elicited by different SARS-CoV-2 vaccines to neutralize VOC replication. Serum is a critical tool for the study of cross-immunity towards naturally occurring SARS-CoV-2 viral variants or model systems such as pseudotyped virus. The availability of reference serum reagents obtained from vaccinated individuals, and produced in volumes large enough to allow distribution, could be used to test for SARS-CoV-2 spike protein binding and neutralization of VOC. This would allow rapid, *in vitro* characterization of the activity of antibodies elicited by different vaccines against emerging VOC. Further, these can serve as a general reference standard to permit comparison of results and enhance interpretation of study outcomes generated via neutralization and/or spike protein binding antibody assays used by different laboratories across the globe.

Efforts to source and generate reagents for continued assessment of SARS-CoV-2 evolution and therapeutic resistance have been championed by the Accelerating COVID-19 Therapeutic Interventions and Vaccines (ACTIV) and the Tracking Resistance and Coronavirus Evolution (TRACE) initiatives. These programs are supported by the National Institutes of Health, the Foundation for the National Institutes of Health, and public-private partnerships. The ACTIV TRACE initiative focuses on the identification of emerging SARS-CoV-2 variants, the evaluation of therapeutic and vaccine resistance, and the assessment of how genetic variation alters viral biology and impacts clinical approaches of disease prevention and treatment. The infrastructure and processes developed by ACTIV TRACE allow for SARS-CoV-2 variant monitoring and data sharing through a five-step approach, which monitors global emergence and circulation of viral mutations, provides cross-reference of sequence data against experimental or clinical viral variant databases, characterizes (both *in vitro* and *in vivo*) VOC using critical-path assays, and shares activity data in a rapid, open manner. An integral part of the ACTIV TRACE working group mission is to assess therapeutic efficacy *in vitro*, generating standardized protocols, reference reagents, and datasets to facilitate greater understanding of VOC impact on vaccine and therapeutic efficacy. Dissemination of SARS-CoV-2 datasets and preclinical assay overviews are freely available through the National Center for Advancing Translation Sciences (NCATS) OpenData Portal: Variant Therapeutic Data Summary (8) while reagents are distributed via collaboration with BEI Resources. The resources generated through the ACTIV TRACE initiative provide a globally accessible platform to further SARS-CoV-2 VOC understanding, prevention, and treatment efforts.

Here, we report the generation of pooled, large volume reference sera reagents obtained from individuals immunized with one of four COVID-19 vaccines. Donors include recipients of the two SARS-CoV-2 mRNA vaccines (BNT162b2, Pfizer and mRNA-1273, Moderna), a replication-incompetent human adenovirus type 26 vaccine (Ad26.COV2.S, Johnson & Johnson), and a recombinant trimeric spike protein expressed by baculovirus and incorporated into a nanoparticle vaccine plus Matrix-M adjuvant (NVX-CoV2373, Novavax). Donors had no evidence of prior infection and passed screening tests required for blood donation. The pools of reference serum have been aliquoted and are available through BEI Resources (https://www.beiresources.org/Home.aspx). We anticipate the creation of additional sera reference standards, based on vaccination and infection convalescence.

## MATERIALS AND METHODS

### Subjects and serum collection

Following written informed consent, 68 individuals participated in the study. The study was approved by the University of Iowa Institutional Review Board (IRB, Committee A). Vaccinated subjects with no history of SARS-CoV-2 infection who lacked detectable antibodies to SARS-CoV-2 nucleocapsid but had antibodies to SARS-CoV-2 spike protein were recruited for a study of T cell responses to SARS-CoV-2. A medical history, whole blood, and peripheral blood mononuclear cells were obtained and processed as described (9). Volunteers who expressed interest were evaluated for blood donation at the Impact Life Blood Services (Coralville, IA). Following a second, written informed consent process, each eligible donor donated one unit (∼ 500 mL) of whole blood, which was collected into a “dry” bag (without anticoagulant). Each unit was allowed to clot at room temperature for ≥ 4 hours, following which serum was recovered into a separate bag without anticoagulant. Each serum product was frozen at ≤ -20°C, and units from each vaccine recipient group were shipped together overnight to BEI Resources on frozen packs. Upon receipt, sera remained partially frozen. The sera were allowed to thaw before being pooled and stored in 1 mL (or 25 mL) labeled tubes. Pooled serum aliquots were frozen and stored at -20°C for subsequent distribution.

### Laboratory methods

Samples from each study volunteer and each pooled serum preparation were evaluated for spike antibody using the LIASON® SARS-CoV-2 S1/S2 IgG assays (Roche Cobas, Roche Diagnostics, Basel, Switzerland) at the University of Iowa Pathology Laboratory and nucleocapsid antibodies using quantitative ELISA (IEQ-CoVN-IgG, IgM, RayBiotech, Peachtree, GA) as recommended by the manufacturer (10, 11). All samples were run in technical replicates. Roche anti-S antibody quantification (U/mL) were converted to estimated World Health Organization IU/mL using the formula recommended by Lukaszuk *et al*. (Roche U/ml / 0.972)(12).

The SARS-CoV-2 spike protein receptor binding domain (RBD) contains a receptor binding motif that interacts with angiotensin-converting enzyme 2 (ACE-2) on the cell surface (13). Antibodies that block cell RBD binding to ACE-2 correlate with neutralizing antibodies (14). Serum inhibition of RBD binding to ACE-2 was measured using a SARS-CoV-2 spike RBD-ACE2 blocking antibody detection ELISA kit (Cell Signaling Technology, Danvars, MA) as recommended by the manufacturer.

## RESULTS

### Study Subject Characteristics

Sixty-eight subjects were evaluated at the Iowa City VA or the University of Iowa. All volunteers were healthy, between 18 and 65 years of age, 43 (63%) were female, 24 male, and 1 was female transgender. The average age of women was 42, and 49 among men. Consistent with local demographics, 2 subjects were African American, 4 were Asian/Pacific Islander, and the remaining 62 were Caucasian, four of whom were Latino. No volunteers had a clinical history of COVID-19. The majority of subjects received BNT162b2 (n=26), followed by mRNA-1273 (n=17), NVX-CoV2373 (n=14, participated in clinical trial), and Ad26.COV2.S (n=9). Two subjects had not received vaccination upon screening.

Thirty-five subjects were excluded from participation in the large volume serum donation study for the following reasons: 18 were not interested in blood donation, 2 had baseline spike and nucleocapsid antibodies detected indicating recent infection, 2 had not received COVID-19 vaccines, 2 were excluded due to failure to meet blood donation screening requirements, 1 was unable to complete donation, and 10 were excluded due to low SARS-CoV-2 spike antibody levels (< 160 WHO IU/mL). The remaining 23 subjects donated 1 unit of whole blood for serum preparation between June 23, 2021 and December 20, 2021. All donors tested negative for Syphilis MHA-TP, HIV-1/2/group O antibody, HIV-1/2-group O RT-PCR, HCV RT-PCR, HCV antibody, HBV RT-PCR, HBsAg, Anti-HB-core antibody, WNV RT-PCR, Zika virus RT-PCR, Babesia RT-PCR if from an endemic state, *Trypanosoma cruzi* antibody, and HTLV-1/2 antibody.

Demographic characteristics of the donors included in each serum pool are shown in Table 1. Six donors received BNT162b2, mRNA-1273, or Ad26.COV2.S vaccines, and 5 donors received NVX-CoV2373. The final volume of each pool was approximately 1.5 L for BNT162b2, mRNA-1273, or Ad26.COV2.S and 1.2 L for NVX-CoV2373. Women accounted for 4 out of 6 of the BNT162b2 and NVX-CoV2373 pools, and 2 out of 6 in the mRNA-1273 pool. One of 5 Ad26.COV2.S donors were female. The age of donors in each pool ranged from 37 to 52, and the timing of the two vaccine doses were similar (range 24 – 29 days, Table 1).

**Table 1.**
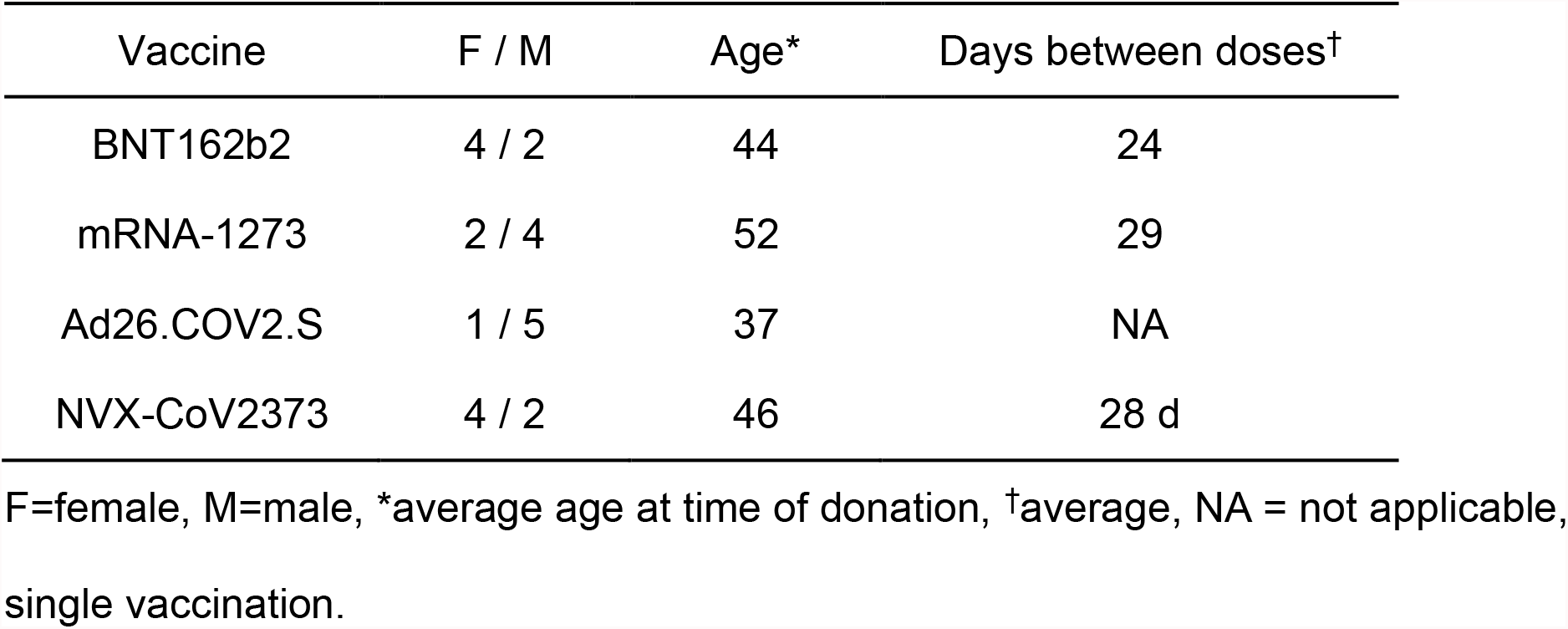

The average number of days following the final vaccination dose varied between vaccines: 69 days for BNT162b2, 102 days for mRNA-1273, and 83 days for Ad26.COV2.S recipients. Since NVX-CoV2373 donors had participated in a double blind, placebo controlled, cross over trial, the time of the vaccination was either January and February 2021 or April and May 2021. Subjects who reported post-vaccination reactions in April or May were estimated to have their second vaccine in May 2021. If the subject had no reaction, or a reaction in January or February, the subjects were estimated to have their second vaccine during their February visit. Based on this, the estimated average time from second NVX-CoV2373 receipt was 152 days (Table 2).

**Table 2.**
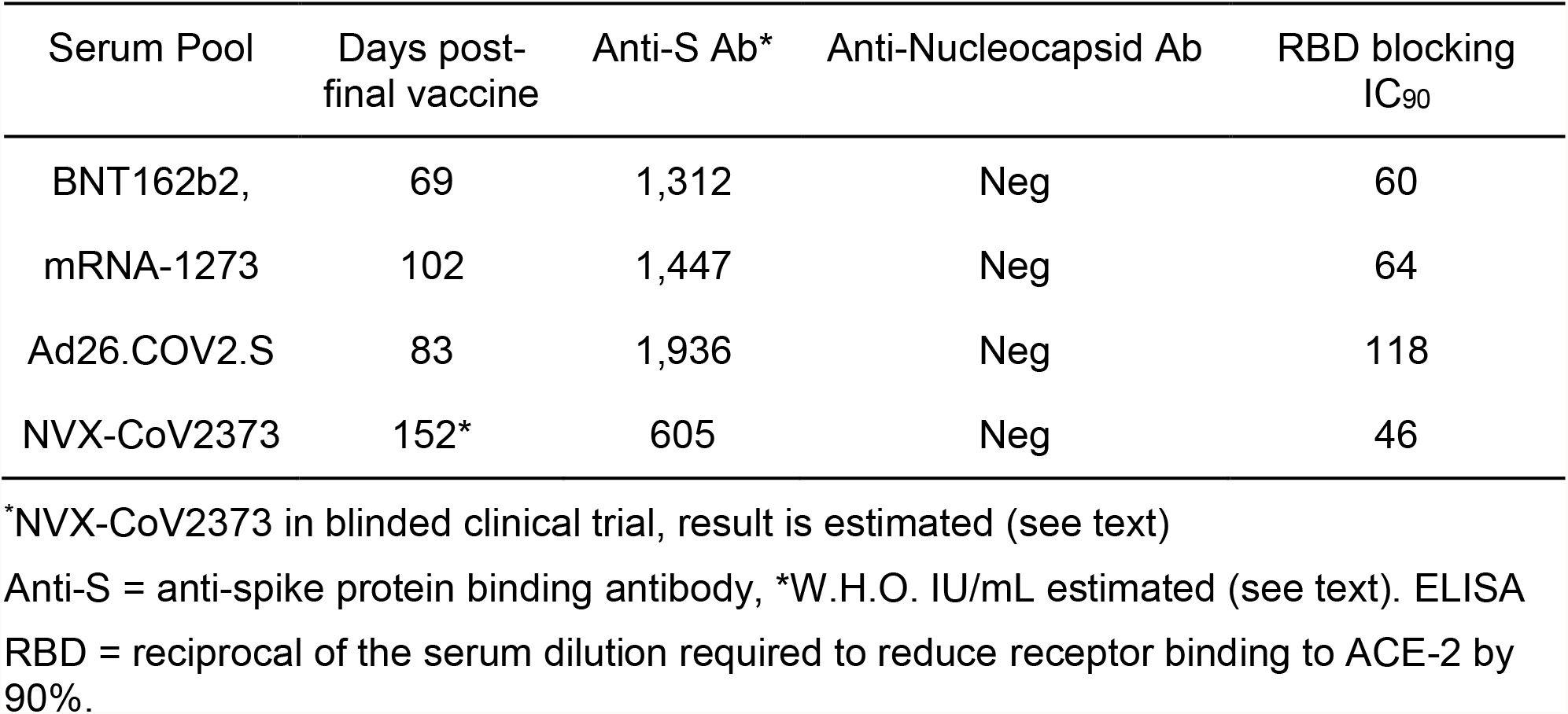

Spike protein binding estimated WHO spike antibody titers ranged from 1276 to 1882 IU/mL for serum pools from BNT162b2, mRNA-1273, and Ad26.COV2.S recipients (Table 2), and failed to correlate with duration from final vaccine receipt. The NVX-CoV2373 recipients had the lowest spike antibody titer (estimated WHO IU/ml of 604, Table 2), which likely reflects the significantly longer time period between final vaccine receipt and serum collection. Nucleoprotein-specific antibodies were not detected in any of the serum pools. Inhibition of RBD binding to ACE-2 reflected total spike antibody concentration (Table 2).

## DISCUSSION

Although post-infection, convalescent sera reference reagents have been generated and made accessible through the National Institute for Biological Standards and Control (NIBSC), to our knowledge the serum pools described herein are the first vaccine-specific reference reagents available for characterizing SARS-CoV-2 spike protein antibodies. Reference reagents are essential for rapid determination of vaccine-specific antibody binding and neutralization of emerging SARS-CoV-2 VOC. The four vaccine-specific sera developed represent three widely used vaccines and a fourth vaccine which has Emergency Use Listing in the European Union and for which an FDA Emergency Use Authorization (EUA) application is thought to be imminent. The large quantity of each reference serum pool should provide continuity for testing current and future SARS-CoV-2 variants and help ascertain if there are differences in binding or function of antibodies generated by different vaccine technologies. Studies are underway to characterize the activity of each serum reference reagent against common VOC.

The SARS-CoV-2 spike protein coding sequence for all four vaccines involved in this study are derived from the same ancestral virus (Wuhan-Hu-1 SARS-CoV-2) with minor differences employed in cloning (reviewed in(15)). Specifically, all four vaccines contain 2 mutations that prevent the conformational change of the more immunogenic pre-fusion spike protein structure into the post-fusion form. Ad26.COV2.S also contains two mutations at the S1/S2 furin cleavage site, which further stabilizes the S protein in the pre-fusion state. The S protein in NVX-CoV2373 has all of the Ad26.COV2.S changes in addition to a deletion in the furin cleavage site. Further, NVX-CoV2373 trimeric (and multi-trimeric) proteins are expressed and purified before formulation as a nanoparticle, which includes a saponin-based adjuvant (Matrix-M™)(15). Due to the high degree of similarity in the spike protein across all four vaccines, differences in antibody responses presumably relate to either host genetic variation, approximate level of vaccine induced immunity/level of wanning immunity post vaccination, and/or differences in vaccine platform (16). Thus, examination of SARS-CoV-2 vaccine-induced antibody binding to their cognate spike protein or neutralization of the Wuhan-Hu-1 strain provides avenues to evaluate different vaccine platforms. In addition, examination of vaccine-specific antibodies binding to emerging VOC spike proteins and the ability of these antibodies to neutralize VOC, allows for an *in vitro* surrogate to aid in predicting vaccine effectiveness Another important aspect of these reference reagents is to enhance the ability to compare results generated by different antibody binding, blocking, or neutralization assays across laboratories around the globe. A plethora of SARS-CoV-2 antibody methods have been developed for diagnostic and research purposes that utilize different platforms (ELISA, lateral flow immunoassays, chemiluminescent immunoassays, SARS-CoV-2 neutralization assays, spike protein pseudotyped neutralization assays, spike protein binding to ACE-2 inhibition, etc.) (17), and it is recognized that results obtained with one assay may vary considerably from that obtained by other laboratories, even when using the same methodology (the so-called reproducibility crisis)(18). There is hope that by making reference serum controls freely available, an improved understanding of the magnitude of the binding or neutralization measured will be gained, and improved harmonization of results between laboratories will emanate.

We foresee, on behalf of the ACTIV TRACE working group, the creation of additional pooled serum reference reagents. These reagents may include samples collected from other vaccination regimens (e.g. subjects who have received vaccine booster doses), or from post-infection convalescent serum with or without vaccination. As new reagents are made available, procedures, protocols, and characterizations will be made available. As a fundamental objective of the ACTIV TRACE initiative, we anticipate widespread distribution of the reagents described in this manuscript via BEI Resources (https://www.beiresources.org/Home.aspx). As reagent-related data are generated, they will be curated, summarized, and shared on the NCATS OpenData Portal (8), thus allowing for the comparison of assays across laboratories and informing on the reproducibility of the global research response to variants of SARS-CoV-2. Development and distribution of these reagents for investigative use against emerging SARS-CoV-2 variants is a driving factor for the ACTIV TRACE working group, as is the timely and open sharing of reagents, protocols, and data to further our understanding and to direct our therapeutic approaches against SARS-CoV-2 VOC as a scientific community.

## Data Availability

All data produced in the present study are availble upon reasonable request to the authors.

## ACKNOWLEGEMENTS

We thank Qing Chang and James McLinden, Ph.D. for helpful discussions, Michelle Rodenburg, Delilah Johnson, A.J. Carr, Susan Herman, Elizabeth Morgan, Deb Pfab, Angel Peguero (University of Iowa Vaccine Evaluation Unit), Barbara Digmann and Tara Dunahoo (Impact Life Blood Services) for assistance with subject recruitment, phlebotomy, and processing. We thank Crystal McClung, Anne Goodling and Gemma Spicka-Proffit at BEI Resources for coordinating shipments of the sera and aliquoting. This work was supported by an intramural program at the National Center for Advancing Translational Sciences (NCATS), National Institute of Allergy and Infectious Diseases, National Institutes of Health, and the HHS Office of the Assistant Secretary for Preparedness and Response (ASPR) Countermeasures Acceleration Group (CAG), the Iowa City Veterans Administration Healthcare System, and by a Merit Review Grant (BX 000207, JTS). The project was funded in part with Federal funds from the National Institute of Allergy and Infectious Diseases, National Institutes of Health, Department of Health and Human Services, under Contract No. HHSN272201600013C, managed by ATCC.

The funders had no role in study design, data collection and interpretation, or the decision to submit the work for publication.

